# ESBL plasmids in *Klebsiella pneumoniae*: diversity, transmission, and contribution to infection burden in the hospital setting

**DOI:** 10.1101/2021.12.20.21268000

**Authors:** Jane Hawkey, Kelly L Wyres, Louise M Judd, Taylor Harshegyi, Luke Blakeway, Ryan R Wick, Adam W J Jenney, Kathryn E Holt

## Abstract

**Background:** Resistance to third-generation cephalosporins, often mediated by extended-spectrum beta-lactamases (ESBLs), is a considerable issue in hospital-associated infections as few drugs remain for treatment. ESBL genes are often located on large plasmids that transfer horizontally between strains and species of Enterobacteriaceae and frequently confer resistance to additional drug classes. While plasmid transmission is recognised to occur in the hospital setting, the frequency and impact of plasmid transmission on infection burden, compared to ESBL+ strain transmission, is not well understood.

**Methods:** We sequenced the genomes of clinical and carriage isolates of *Klebsiella pneumoniae* species complex from a year-long hospital surveillance study to investigate ESBL burden and plasmid transmission in an Australian hospital. Long-term persistence of a key transmitted ESBL+ plasmid was investigated via sequencing of ceftriaxone-resistant isolates during four years of follow-up, beginning three years after the initial study.

**Results:** We found 25 distinct ESBL plasmids. One (Plasmid A, carrying *bla*_CTX-M-15_ in an IncF backbone similar to pKPN-307) was transmitted at least four times into different *Klebsiella* species/lineages and was responsible for half of all ESBL episodes during the initial one-year study period. Three of the Plasmid A-positive strains persisted locally 3–6 years later, and Plasmid A was detected in two additional strain backgrounds. Overall Plasmid A accounted for 21% of ESBL+ infections in the follow-up period.

**Conclusions:** Whilst ESBL plasmid transmission events were rare in this setting, they had a significant and sustained impact on the burden of ceftriaxone-resistant and multidrug-resistant infections.

**summary:** We detected a *bla*_CTX-M-15_ plasmid (Plasmid A) that transferred four times into different *Klebsiella* lineages, causing 50% of ESBL episodes during the initial study. Three Plasmid A-positive strains persisted locally 3–6 years later, accounting for 21% of ESBL+ infections.

## Introduction

Healthcare-associated infections (HAIs) are a top global health priority. In industrialised countries, the HAI burden is greater than that of all other communicable diseases combined [1]. Antimicrobial resistant (AMR) HAIs are particularly problematic as they reduce treatment options, and the World Health Organisation recognises third-generation cephalosporin-resistant (3GCR) and carbapenem-resistant Gram-negative bacterial HAI pathogens, including *Klebsiella pneumoniae* and *Escherichia coli*, as a critical threat [2]. These resistance phenotypes are typically mediated by horizontally acquired extended-spectrum beta-lactamase (ESBL) and carbapenemase genes, encoded on large plasmids that circulate amongst different species. As a result, transmission of AMR HAIs within healthcare settings can result from transmission of a resistant strain, or of a resistance-encoding plasmid between strains of the same or different species [3]. *K. pneumoniae* is a well-known reservoir of AMR plasmids, recognised for its ability to acquire and transfer these plasmids across its population and into other species [4,5].

Genomics is a powerful approach for investigating transmission of AMR HAIs, which is gradually being implemented to detect and contain transmission of AMR pathogens [6–8]. Lengthy hospital outbreaks (3-6 years) have been documented due to plasmid transmission (frequently involving *Klebsiella*) [9–11], and metagenomic studies show that AMR plasmids are ubiquitous in hospitals [12]. However, genomics surveillance studies typically focus on identifying strain transmission rather than plasmid transmission, and the contribution of the latter to HAI burden is poorly understood.

Here we examine the diversity and transmission of ESBL plasmids in *K. pneumoniae* species complex (KpSC) isolates collected during a one-year surveillance period in a large tertiary hospital and referral network in Melbourne, Australia [13–15], and explore the contribution of ESBL plasmid transmission to infection burden in both the short term (year-long study period) and longer term (4 years of follow-up, beginning 3 years after the primary study).

## Methods

### Ethical approval

The primary study was approved by the Alfred Health Human Research Ethics Committee (AHHREC), Project numbers #550/12 (19 February 2013) and #526/13 (10 December 2013). Analysis of clinical isolates 2017-2020 was approved by AHHREC, Project number #371/19 (2 July 2019).

### Specimen collection

The primary KpSC collection comprised 440 *Klebsiella pneumoniae, Klebsiella variicola* and *Klebsiella quasipneumoniae* isolated at the Alfred Hospital Microbiological Diagnostic Laboratory from patients at hospitals in the Alfred Health network, Melbourne, during a prospective surveillance study (KASPAH) between April 2013 and March 2014 (**Supplementary Table 1**). KASPAH study protocols are reported elsewhere [13–15]. Briefly, gut carriage (colonising) isolates (n=108) were cultured from rectal screening swabs in either the Alfred Hospital ICU (33% of patients screened) [13] or Caulfield Hospital geriatric wards (31% screened) [14]. Infection isolates (n=332) represent all KpSC clinical isolates identified by standard diagnostic protocols across the study period [15]. In January to March 2014, 3GCR Gram-negative carriage and infection isolates were also collected from Alfred Hospital ICU patients as described previously [16], and those identified as Enterobacteriaceae were included here (n=74). All isolates were subjected to antimicrobial susceptibility testing using Vitek2 GN AST cards (bioMérieux) and interpreted using EUCAST cut-offs (2020).

To determine whether KpSC lineages harbouring a plasmid of interest (Plasmid A) during the primary study period persisted 3-6 years later, we interrogated all 3GCR KpSC clinical isolates identified by the same diagnostic laboratory between March 2017 and December 2020 (n=450, identified as *K. pneumoniae* or *K. variicola* by MALDI-TOF (Bruker) and resistant to ceftriaxone (MIC ≥2 mg/mL)).

### Whole-genome sequence (WGS) analysis

DNA was extracted and sequenced using the Illumina platforms as previously reported [13–16]. A subset (n=70) of ESBL+ isolates from the KASPAH study were selected for additional sequencing via the Oxford Nanopore Technologies MinION platform as described previously [17]. These included at least one representative of each unique combination of multi-locus sequence type (ST) and ESBL gene. Representatives of *bla*_CTX-M-15_ positive KpSC STs identified in the 2017-2020 follow-up period were also subjected to MinION sequencing. Genomes were assembled using Unicycler v0.4.7 [18] and annotated with Prokka v1.14 [19]. Completed genome assemblies were deposited in GenBank and Illumina read sets in the NCBI Sequence Read Archive (accessions in **Supplementary Table 1**). STs were identified for each genome using Kleborate v1.1 [20] for KpSC, and mlst v2.19 (https://github.com/tseemann/mlst) for *Escherichia*. AMR genes were detected using Kleborate v1.1 [20].

### Identification and comparison of ESBL/carbapenemase plasmids

For each ESBL or carbapenemase gene identified in multiple genomes, the plasmid sequences harbouring them were compared pairwise using two similarity metrics: (i) Mash similarity (1–[Mash distance], calculated using Mash v2.1.1 [21]), and (ii) gene content Jaccard similarity ([genes in common]/[total genes in either plasmid]). Pairs of plasmids with Mash similarity ≥0.98 and gene content similarity ≥0.8 were designated the same plasmid (thresholds determined empirically, see **Supplementary Figure 1**). Plasmid replicon markers and MOB genes were detected using Mobtyper v1.4.9 [22], and copy numbers estimated by Unicycler.

For n=418 genomes in the 2017-2020 follow-up collection, we used read mapping to determine the presence of Plasmid A, represented by pINF329. Illumina reads were mapped to pINF329 (accession LR890241) using RedDog (https://github.com/scwatts/reddog-nf) to determine coverage and depth of the plasmid sequence as described previously [23]. Genomes were considered Plasmid A-positive if they had ≥80% mapping coverage and <10 single nucleotide variants (SNVs) compared to the pINF329 reference sequence (n=133 genomes).

### Phylogenetic analyses

We generated core-genome phylogenies for Plasmid A, and chromosomes of *K. pneumoniae* ST323 (n=40), ST29 (n=17), ST2856 (n=10) and *K. variicola* ST347 (n=54). Illumina reads were mapped to completed reference sequences generated from study isolates (pINF329 for Plasmid A, accession LR890241; INF018 for ST323, accession LR890493; INF250 for ST29, accession LR890374; KPN342 for ST2856, accession CP089384; INF345 for ST347, accession LR890399) using RedDog to identify SNVs as described above. For additional context, chromosome SNV alignments were supplemented with genomes of the same ST from public isolate collections [20]. Phylogenies were inferred using RAxML v8.2.9 [24] with a general time-reversible substitution model. See **Supplementary Methods** for further details.

## Results

Year-long surveillance of KpSC clinical infection (hospital-wide) and carriage (ICU only) identified infections in 303 patients and colonisation in 102 (including 14 with clinical infection) [13,15]. WGS identified ESBLs in 47 (15%) infections and 10 (10%) carriage episodes [15]. Additional screening for ESBL+ Enterobacteriaceae in ICU patients during the last three months of the study identified 25 non-KpSC isolates for comparison (n=24 *Escherichia coli*, n=1 *Escherichia marmotae*) representing 6 infection and 19 rectal carriage episodes [16].

All ESBL+ isolates were confirmed 3GCR, displaying ceftriaxone MIC >2 mg/L. Seven different acquired ESBL genes were detected in the KpSC isolates; *bla*_CTX-M-15_ was the most common, present in 85% of all ESBL+ genomes (n=62) (**Figure 1a**). All KpSC genomes carrying *bla*_VEB-1_ also carried *bla*_CTX-M-15_. Six acquired ESBL genes were detected in the *Escherichia* isolates, again bla_CTX-M-15_ was the most common (n=9, 37% of ESBL+ *Escherichia*) (**Figure 1b**). Twelve KpSC isolates (all ESBL+) carried carbapenemase genes: *bla*_OXA-48_ (n=8 genomes, five episodes, all *K. pneumoniae* ST231 and also carrying *bla*_CTX-M-15_) and *bla*_IMP-4_ (n=4 genomes, two episodes, all *K. pneumoniae* ST340 and also carrying *bla*_CTX-M-15_). ESBL+ isolates harboured a significantly higher burden of acquired AMR genes than ESBL-ones, amongst both KpSC (median 12 vs 0, p<10^−15^ using Wilcoxon rank sum test, one representative per episode) and *Escherichia* (median 10 vs 2, Wilcoxon test, p=5.4×10^−5^) (**Figure 2a**).

**Figure 1:**
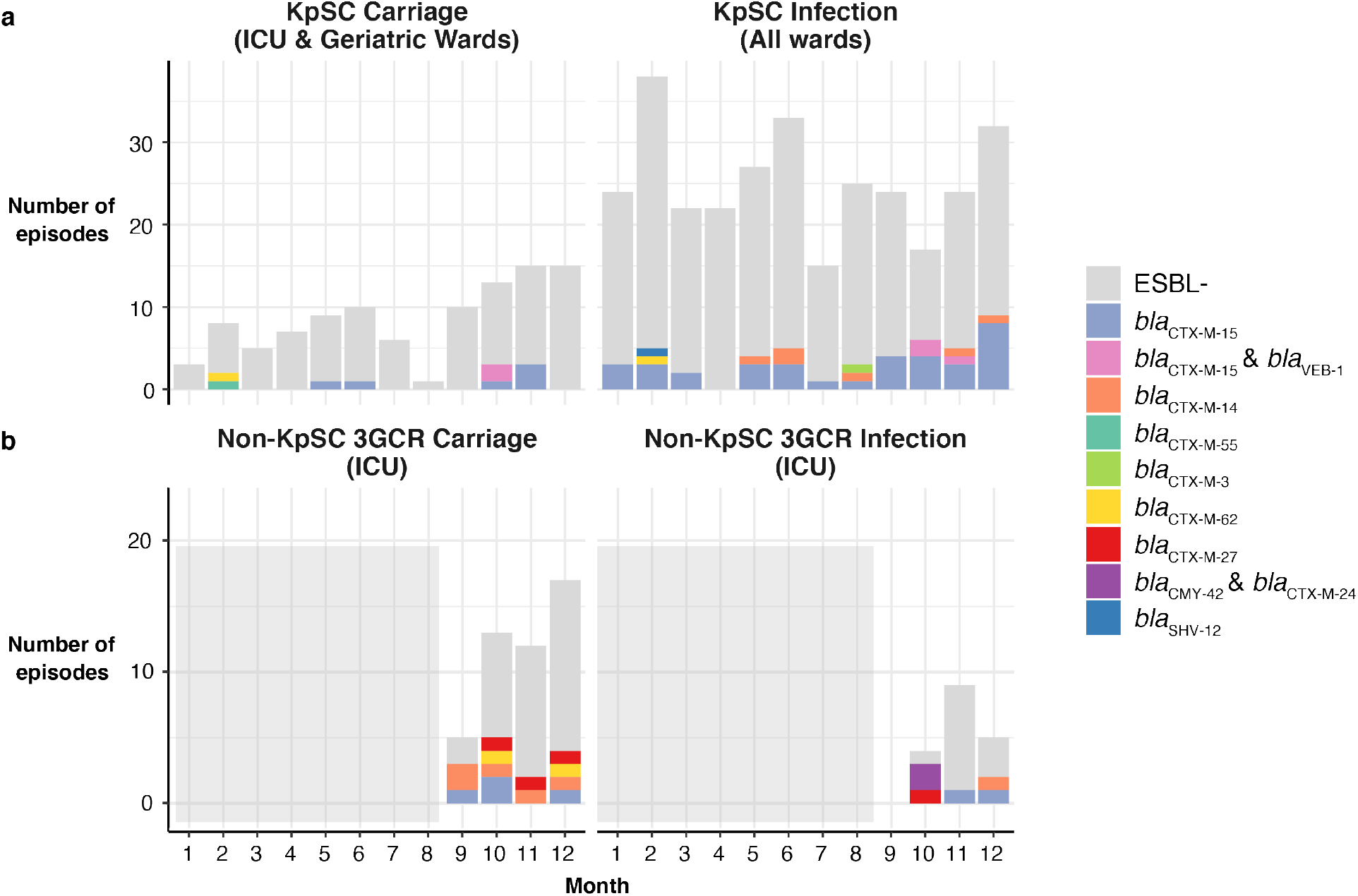
Number of carriage and infection episodes detected per month during the year-long KASPAH surveillance study. **a**, KpSC isolates (regardless of 3GCR status); **b**, 3GCR non-KpSC Enterobacteriaceae (collected in ICU and during the final three months only). Bars are coloured by acquired ESBL gene, as per legend. Light grey boxes in panel **b** indicate no data available for that time period.

**Figure 2:**
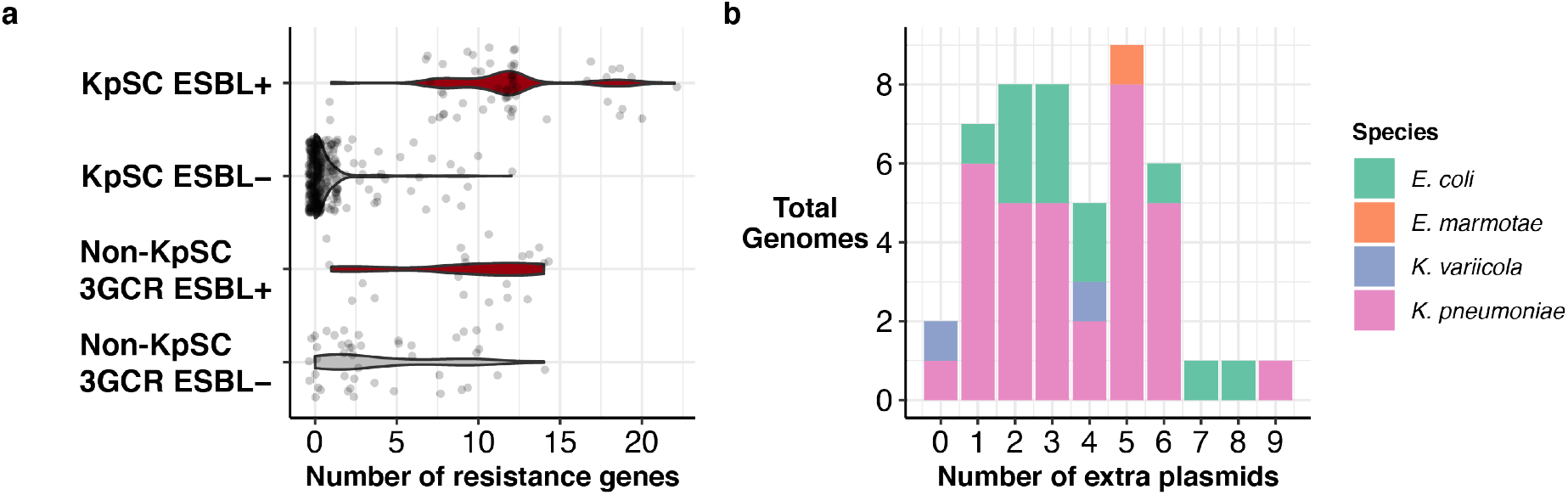
Acquired AMR gene burden and plasmid burden. **a**, Distribution of acquired AMR genes across isolates collected in the year-long KASPAH study, stratified by sample group (KpSC, capturing both 3GCR and susceptible isolates; 3GCR, capturing only 3GCR isolates of non-*Klebsiella* Enterobacteriaceae species) and acquired ESBL status. Violin plots are coloured by ESBL status (ESBL+, red; ESBL-, grey). **b**, Histogram showing the distribution of non-ESBL plasmid counts for ESBL plasmid-positive genomes. Bars are coloured by species as per legend.

### ESBL plasmid diversity

To determine the genetic context of ESBL genes, we generated complete closed reference sequences for n=67/104 ESBL+ genomes (64%), including at least one for each unique combination of ESBL gene and species/ST, representing n=48/77 of all ESBL episodes (62%). Most of the completed KpSC genomes (n=48/52, 92%) carried a plasmid-borne ESBL gene (including n=14 with an additional chromosomal copy); the rest (n=4/52, 8%) carried a chromosomal ESBL gene only (*bla*_CTX-M-15_). Similarly, amongst *Escherichia* genomes, 87% (n=13/15) carried a plasmid-borne gene (including n=2 with an additional chromosomal copy) and 13% (n=2/15) carried a chromosomal ESBL gene only (*bla*_CTX-M-15_).

Using pairwise similarity scores to compare all 61 completed ESBL plasmid sequences (see **Methods**) we identified 25 distinct plasmids. Those found in only a single genome were labelled with their host isolate, and those found in ≥2 genomes were labelled Plasmid A, Plasmid B etc. Each ESBL plasmid was restricted to either KpSC (n=12) or *Escherichia* (n=13) (**Table 1**). The majority (n=17, 68%) were IncF type plasmids, ranging in size from 45–245 kbp with estimated copy number 1.3–3 per cell, and accounting for 70% of the *bla*_CTX-M_ genes that were identified (**Table 1, Supplementary Table 2**). Other notable ESBL plasmids included two IncN plasmids carrying *bla*_CTX-M-62_ (both copy number ∼5x; one 59 kbp, the other 102 kbp and also harbouring an IncR replicon marker), and a small (10 kbp) high copy number (16x) plasmid carrying *bla*_CTX-M-14_ in *E. coli* ST48 (**Table 1, Supplementary Table 2**).

**Table 1:** Unique ESBL plasmids found in this study. **#**Pt, number of patients; #G, number of genomes (completed & draft). Kp – *K. pneumoniae*; Kv – *K. variicola*; Ec – *E. coli*; Em – *E. marmotae*.

| Plasmid | ESBL | #Pt | #G | Organisms | Median size (bp) | Inc type(s) | MOB | Median copy number | # AMR classes (genes) |
| --- | --- | --- | --- | --- | --- | --- | --- | --- | --- |
| ***KpSC ESBL Plasmids*** |  |  |  |  |  |  |  |  |  |
| Plasmid A | *bla*<sub>CTX-M-15</sub> | 18 | 39 | Kp (4 STs)<br>Kv ST347 | 243,577 | FIB, FII | F | 1.30 | 3-5 (5-8) |
| Plasmid B | *bla*<sub>CTX-M-15</sub><br>*bla*<sub>VEB-1</sub> | 4 | 6 | Kp ST231 | 128,238 | C (ST3) | H | 1.49 | 6 (12) |
| Plasmid C | *bla*<sub>CTX-M-15</sub> | 4 | 4 | Kp ST491 | 190,324 | FIA, FII | F | 1.49 | 6 (6) |
| Plasmid D | *bla*<sub>CTX-M-15</sub> | 3 | 8 | Kp ST340 | 89,345 | FIA, R | - | 2.20 | 5 (9) |
| pINF044 | *bla*<sub>CTX-M-15</sub> | 1 | 1 | Kp ST437 | 227,214 | FIB, FII | F | 1.07 | 6 (9) |
| pINF161 | *bla*<sub>CTX-M-14</sub> | 1 | 1 | Kp ST661 | 89,062 | FIIA | P | 1.15 | 0 (0) |
| Plasmid E | *bla*<sub>CTX-M-14</sub> | 3 | 3 | Kp ST17 | 214,317 | FIB, FII | F | 1.24 | 7 (10-12) |
| Plasmid F | *bla*<sub>CTX-M-14</sub> | 2 | 2 | Kp ST3062 | 159,592 | FIB,<br>FIIA, FII,<br>FIA | F | 2.58 | 5 (8) |
| Plasmid G | *bla*<sub>CTX-M-62</sub> | 2 | 2 | Kp ST15 | 102,016 | N, R | P, F | 5.97 | 8 (11) |
| pINF223 | *bla*<sub>CTX-M-3</sub> | 1 | 1 | Kp ST3072 | 60,529 | FII | P | 2.98 | 2 (2) |
| pKSB1_1B | *bla*<sub>CTX-M-55</sub> | 1 | 1 | Kp ST1213 | 113,743 | I2 | P | 1.30 | 0 (0) |
| pINF058 | *bla*<sub>SHV-12</sub> | 1 | 1 | Kv ST3076 | 279,735 | H | H | 1.55 | 8 (8) |
| ***Escherichia ESBL plasmids*** |  |  |  |  |  |  |  |  |  |
| pMSB1_1A | *bla*<sub>CTX-M-15</sub> | 1 | 1 | Ec ST617 | 173,787 | FIB,<br>FIIA, FII,<br>FIA | F | 1.53 | 7 (10) |
| pMSB1_9I | *bla*<sub>CTX-M-15</sub> | 1 | 1 | Ec ST10 | 44,618 | FIIA, FII | F | 1.30 | 0 (0) |
| pMINF_8D | *bla*<sub>CTX-M-15</sub> | 1 | 1 | Ec ST10 | 115,278 | unknown | none | 1.72 | 0 (0) |
| pMINF_1D | *bla*<sub>CTX-M-15</sub> | 1 | 1 | Ec ST141 | 125,802 | FIB,<br>FIIA, FII,<br>FIA | F | 1.55 | 1 (1) |
| Plasmid H | *bla*<sub>CTX-M-14</sub> | 2 | 2 | Ec ST48 | 10,057 | none | none | 15.81 | 2 (2) |
| pMINF_2E | *bla*<sub>CTX-M-14</sub> | 1 | 1 | Ec ST602 | 207,369 | FIB,<br>FIIA, FII | P, F | 1.58 | 0 (0) |
| Plasmid I | *bla*<sub>CTX-M-14</sub> | 2 | 2 | Ec ST38 | 70,350 | FIIA, FII | F | 2.71 | 0 (0) |
| pMSB1_5C | *bla*<sub>CTX-M-14</sub> | 1 | 1 | Em ST3727 | 70,361 | FIIA, FII | F | 2.63 | 0 (0) |
| Plasmid J | *bla*<sub>CTX-M-27</sub> | 2 | 3 | Ec ST131 | 112,822 | FIB,<br>FIIA, FII,<br>FIA | F | 1.95 | 0 (0) |
| Plasmid K | *bla*<sub>CTX-M-27</sub> | 1 | 2 | Ec ST38 | 160,498 | FIB,<br>FIIA, FII,<br>FIA | F | 1.91 | 6 (9) |
| pMINF_9A | *bla*<sub>CTX-M-27</sub> | 1 | 1 | Ec ST648 | 139,312 | FIB,<br>FIIA, FII,<br>FIA | F | 1.79 | 5 (7) |
| Plasmid L | *bla*<sub>CTX-M-62</sub> | 2 | 2 | Ec ST176 | 59,604 | N | F | 4.83 | 5 (5) |
| Plasmid O | *bla*<sub>CMY-42</sub> | 2 | 2 | Ec ST354 | 47,582 | I1 | P | 2.05 | 0 (0) |

Ten of 12 KpSC ESBL plasmids carried additional AMR genes (median 8.5 AMR genes per plasmid); however, only half (n=6/13) of the *Escherichia* ESBL+ plasmids carried additional AMR genes (median 6 per plasmid) (**Table 1, Supplementary Table 2**). Nearly all (n=45/48, 93%) of the completed ESBL plasmid-carrying KpSC genomes carried at least one additional plasmid (range 0-9 plasmids), and 70% carried at least three (**Figure 2b**). The majority of additional plasmids did not carry AMR genes, except for two STs carrying carbapenemase plasmids: *K. pneumoniae* ST340 with *bla*_IMP-4_ (n=4 isolates) and *K. pneumoniae* ST231 with *bla*_OXA-48_ (n=8 isolates) (**Supplementary Table 2, Supplementary Text**). All of the completed ESBL plasmid-carrying *E. coli* genomes carried at least one additional plasmid (range 1-8) (**Figure 2b**), the majority of which lacked AMR genes (n=8/13, 62%).

### Plasmid transmission network

We used the completed genomes to investigate the evidence for nosocomial transmission of ESBL plasmids between distinct KpSC strain backgrounds (defined by ST), by constructing a network in which nodes represent ESBL+ plasmids identified in KpSC (**Figure 3**). The vast majority of unique ESBL plasmids were detected in either a single genome (n=13/25) or a small number of genomes of a single ST (n=11/25, 2-8 genomes each) (**Table 1, Figure 3**). However, Plasmid A (which carried *bla*_CTX-M-15_) was identified in 41 isolates belonging to four *K. pneumoniae* STs (ST323, ST29, ST5822, ST221) and *K. variicola* (ST347, see **Figure 3**). Plasmid A was highly conserved between isolates, with median pairwise gene content similarity 0.93 (range 0.84-1) and median Mash distance 0.998 (range 0.996-1; see **Figure 3, Supplementary Table 2**). Median 0 SNVs were detected amongst Plasmid A sequences (19 identical, range 0-54 SNVs), regardless of host chromosomal ST, with only occasional differences in plasmid gene content (**Supplementary Figure 2**). Taken together, these data support transmission of Plasmid A between multiple *K. pneumoniae* lineages and one *K. variicola* lineage.

**Figure 3:**
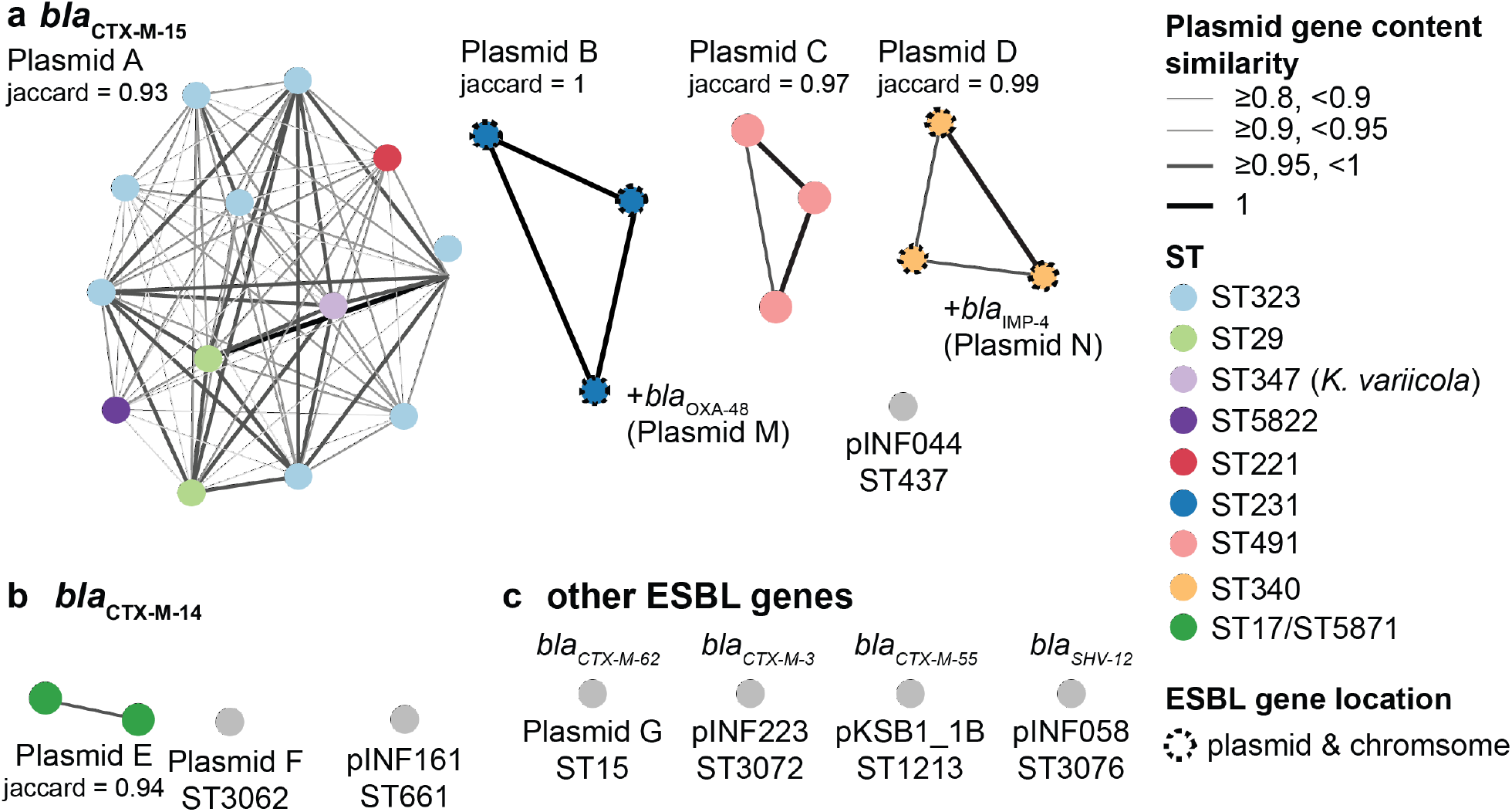
ESBL plasmid cluster network for all completed KpSC ESBL+ genomes. **a**, *bla*_CTX-M-15_ plasmids; **b**, *bla*_CTX-M-14_ plasmids; **c**, other detected ESBLs. Each node represents a completed genome (from one representative isolate per patient), coloured by the bacterial host cell ST as per legend. Dashed outline surrounding dot indicates if the ESBL is carried on both a plasmid and the chromosome. Nodes are connected if they share a similar plasmid sequence, defined as mash similarity ≥0.98 and gene content similarity ≥0.8 (see **Supplementary Figure 1**); line colour and width indicate plasmid gene content similarity (as per legend). Clusters comprising ≥2 genomes are labelled with the median jaccard gene similarity score. ‘+*bla*_OXA-48_ (Plasmid M)’ and ‘+*bla*_IMP-4_ (Plasmid N)’ indicate STs that carry an additional carbapenemase plasmid (details in **Supplementary Table 2**).

Applying a similar analysis to the carbapenemase plasmids revealed an IncL plasmid (Plasmid M carrying bla_OXA-48_, **Supplementary Table 2**) in two different host strains (*E. coli* ST38 and *K. pneumoniae* ST231) from the same patient. The sample collection dates were consistent with transfer of Plasmid M from *K. pneumoniae* ST231 to *E. coli* ST38 in this patient’s gut (**Supplementary Figure 3, Supplementary Text**).

### Dissemination of Plasmid A and contribution to ESBL burden

Overall Plasmid A was present in 51% (n=29/57) of KpSC ESBL+ episodes (53% of ESBL+ infections) during the study period. The remaining 49% (n=28/57) of KpSC ESBL+ episodes were attributed to plasmids/strains isolated from one patient only (**Figure 5a**). *K. pneumoniae* ST323 was the most common host strain for Plasmid A (n=22). Plasmid A-positive ST323 were detected throughout the entire study period and accounted for 29% of ESBL+ KpSC episodes (25.5% of infections, 50% of carriage) (**Figure 4**). Plasmid A-positive ST323 isolates from distinct patients differed from each other by 0-55 chromosomal SNVs (median 22, see **Supplementary Figure 4**), consistent with clonal transmission of this strain within the hospital network as previously reported [14].

**Figure 4:**
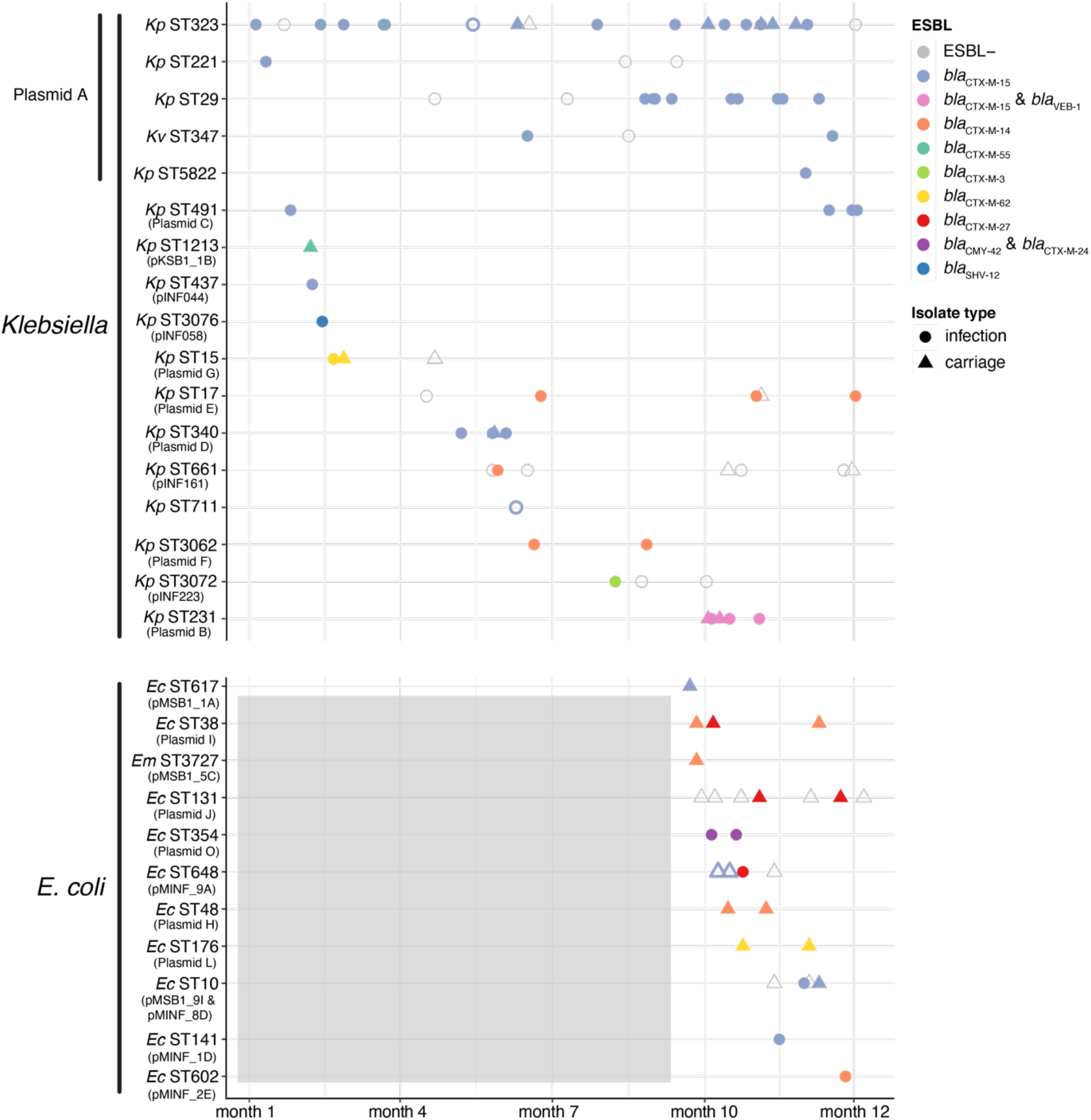
Timeline of ESBL+ infection and carriage episodes. Each episode is represented by a single isolate, grouped in rows by the isolate ST. Colours indicate the ESBL enzyme encoded in the genome (as per legend), and shapes indicate whether the episode was carriage (circle) or infection (triangle). Presence of the ESBL plasmid most commonly associated with the ST is indicated by filled shapes (with plasmid name listed under ST on the y-axis). ESBL genes on the chromosome rather than plasmid are indicated by shapes with no fill and a coloured outline. Grey box indicates that no data was available for this period for the *Escherichia* STs. Kp – *K. pneumoniae*; Kv – *K. variicola*; Ec – *E. coli*; Em – *E. marmotae*.

**Figure 5:**
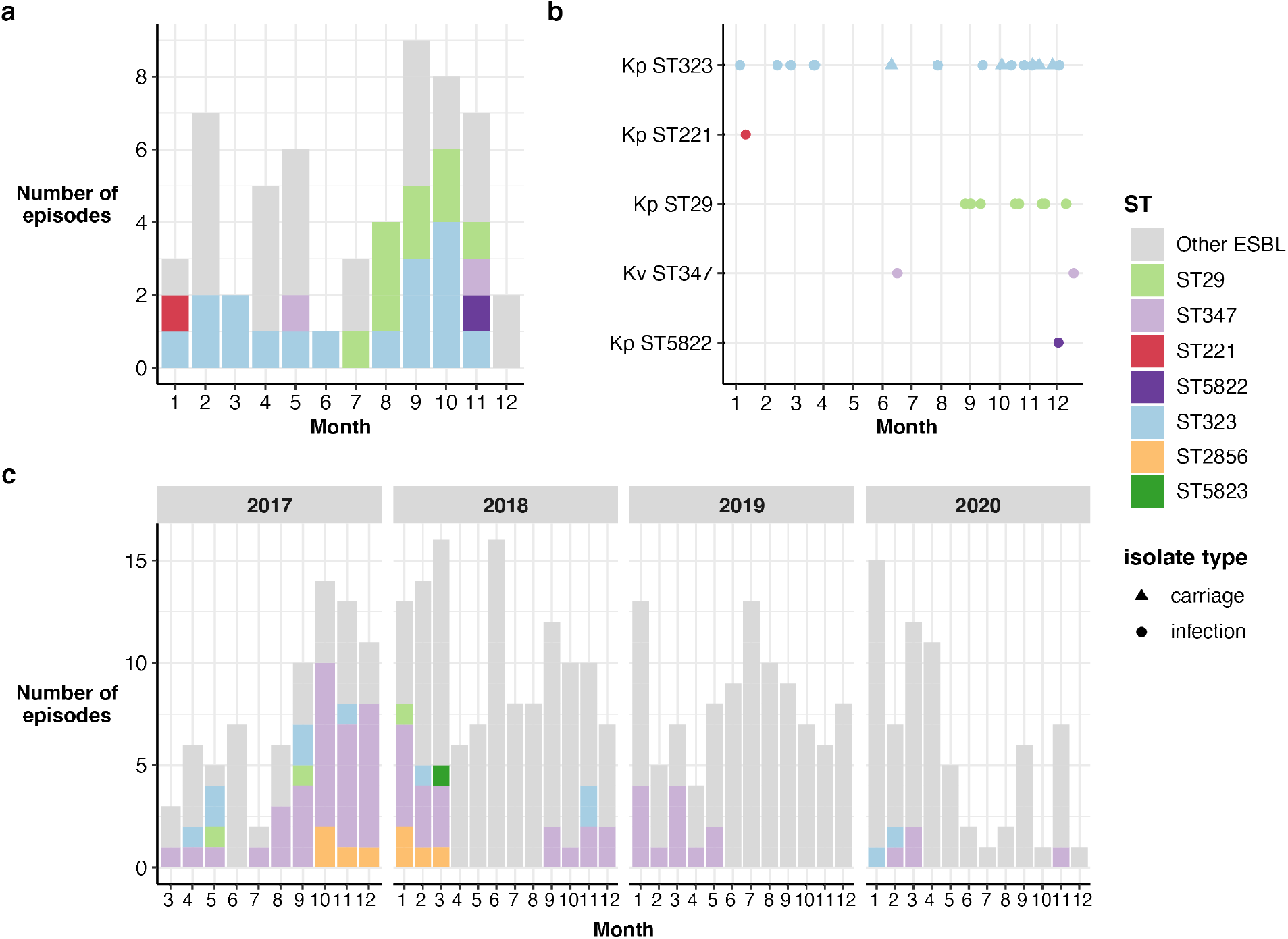
Presence of Plasmid A over time. **a**, Bar height indicates total number of ESBL episodes across the initial year-long study period, by month. Bars are coloured by Plasmid A positive ST episodes as per legend, and grey for non-Plasmid A ESBL episodes. **b**, Timeline of Plasmid A positive episodes during the initial year-long study period. Each point represents an episode, with colour indicating ST (as per legend) and shape indicating whether the episode was infection (circle) or carriage (triangle). **c**, Bar height indicates the total number of 3GCR KpSC episodes per month from March 2017 – December 2020. Bars are coloured by ST as per legend if the episode was positive for Plasmid A.

We hypothesise that Plasmid A was originally hosted in ST323 and subsequently transmitted into four other KpSC STs. This is supported by identification of an outgroup ST323 genome (EuSCAPE_IT380, accession ERR1217451, isolated in Italy in 2013 [25]) carrying Plasmid A (2 plasmid SNVs, 59 chromosomal SNVs, **Supplementary Figure 4**). Additionally, two ST29 isolates collected in months 4 and 7 did not carry Plasmid A (INF206 and INF122, **Figure 4**), and were evolutionarily distant from the 11 Plasmid A-positive ST29 genomes recovered in months 8-12 (INF206=10,341 core genome SNVs, INF122=106 SNVs; **Supplementary Figure 4**) [14]. Similarly, a Plasmid A-negative *K. variicola* ST347 isolate collected in month 8 (INF238, **Figure 4**) was distant to subsequent ST347 isolates (35,020 core genome SNVs; **Supplementary Figure 4**). Analysis of the Plasmid A backbone sequence showed that the variants present in ST29, ST5822, ST221 and ST347 genomes were clustered in a single sub-clade of the Plasmid A phylogeny, whereas ST323 variants were more diverse and present both within and outside of this sub-clade (**Supplementary Figure 2**).

We detected only a single episode each of ST221 and ST5822 with Plasmid A, but the data supported nosocomial transmission of the ST29 and ST347 strains following acquisition of Plasmid A (median chromosomal SNVs between isolates from different patients = 1 for both STs, range 0-5). Assuming all four STs acquired Plasmid A from ST323, these plasmid transmission events and subsequent onward transmission of the resulting ESBL+ strains would account for 23% (n=13/57) of KpSC all ESBL+ episodes (28% of ESBL+ infections) during the one-year study period, including 54% of those in the last 3 months (**Figure 5a,b**).

### Long-term persistence of Plasmid A in diverse strain backgrounds

We screened for Plasmid A in additional WGS data from 3GCR KpSC clinical isolates collected 3-6 years later from the same hospital laboratory (n=418, representing 92% of all 3GCR KpSC clinical isolates during March 2017 – Dec 2020). These 418 isolates represented 365 infection episodes (57% urinary tract, 13% bacteraemia, 12% respiratory, 5.8% wound/soft tissue). Plasmid A was identified in 35% of all sequenced 3GCR isolates and 21.2% of all 3GCR infection episodes (24%, 21%, 42% and 8.5% of 3GCR urinary, bacteraemia, wound/soft tissue, and respiratory episodes, respectively). We observed the same three expanded STs seen in the original study (n=11 ST323, n=3 ST29, n=69 ST347), as well as two additional STs (n=11 ST2856, n=1 ST5823) (**Figure 5c**). Phylogenetic analysis of the host chromosomes was consistent with persistence of the plasmid during long-term local clonal expansion of the ST323, ST29 and ST347 host strains (**Supplementary Figure 4**). Inspection of completed genome sequences for one genome per ST (KPN029, KPN110, KPN392, KPN342, KPN692) confirmed that the more recent isolates carried Plasmid A, with few structural changes (**Supplementary Figure 5a**). Plasmid A was most prevalent from March 2017 to March 2018, accounting for 52% of 3GCR episodes (n=62/120, **Figure 5c**). From April 2018 onwards, Plasmid A episodes were rare (<10%), except for a cluster of infections between September 2018 and May 2019 during which time Plasmid A-positive ST347 accounted for 27.5% of 3GCR infections (**Figure 5c**).

## Discussion

Here, we present a systematic comparison of ESBL strain and plasmid transmission over a year in a single hospital network. Whilst there was no evidence of plasmid transmission for most ESBL+ plasmids, Plasmid A was the exception and we speculate that ST323 was the original donor strain. Whilst we cannot be certain that Plasmid A transmission events occurred within our hospital (ST323 has been detected in other Melbourne hospitals [8]), it is most likely that they occurred in a hospital setting under selective pressure from antibiotics, since ESBL KpSC is known to be relatively rare outside of the hospital environment in Australia and other high-income countries [26,27].

Our results indicate that Plasmid A is stable in at least some strain backgrounds, with three out of six putative recipient strains going on to spread in the hospital in parallel with the dissemination of Plasmid A-positive ST323. Compared with the other ESBL+ plasmids identified in this study, which showed no evidence of transfer between strains, Plasmid A likely has properties that (i) make it efficient at horizontal transfer, and/or (ii) reduce the fitness cost and thus facilitate its stable maintenance. Plasmid A is very similar to the IncFIB/IncFII *bla*_CTX-M-15_ plasmid found in the globally disseminated *K. pneumoniae* clone ST307 (pKPN-307, median Mash distance 0.99, 70% gene similarity to Plasmid A, with some differences in gene order; **Supplementary Figure 5b**), which has been stably maintained for decades [28]. pKPN-307 carries five distinct virulence clusters that are hypothesised to aid long term survival of ST307 outside of the human host [29]; all five of these clusters are conserved in Plasmid A, in addition to heavy metal resistance operons (**Supplementary Figure 5c**). Other IncFIB/IncFII plasmids detected in the globally distributed *K. pneumoniae* ST258 also carry many of these genes [30], suggesting that they may be a feature of this plasmid background. Understanding what plasmid and/or host strain properties enable stable maintenance of large AMR plasmids within a particular genetic background is vital to improving our understanding of how AMR strains emerge and persist [31].

ESBL infections contribute to increased length of hospital stays, resulting in higher healthcare costs [32]. Models examining genomic surveillance have been shown to reduce overall healthcare costs [33], but have not considered the impact of plasmid transmission on AMR burden. Here, prevention of onward transmission of ST323 would have reduced the opportunity for Plasmid A to spread and create novel ESBL+ strains. This could potentially have prevented half of all ESBL episodes in the primary one-year study period, and one-fifth of all 3GCR episodes in the follow-up period 3-6 years later. Whilst our study focused on ESBL plasmids, those carrying carbapenemases transfer via the same mechanisms and show similar transmission dynamics. Like ESBL plasmids, some carbapenemase plasmids have been found to transmit amongst the Enterobacteriaceae in hospital environments [34,35], and in particular *Klebsiella* has been known to harbour multiple ESBL and carbapenemase plasmids in a single cell [36,37]. This study highlights the need for improved approaches to detect plasmid transmission using genomic data within an infection control framework.

## Data Availability

All accession numbers are provided in Supplementary Table 1.

