## Supplementary figures and images for "ESBL plasmids in *Klebsiella pneumoniae*: diversity, transmission, and contribution to infection burden in the hospital setting"

### Supplementary Figure 1

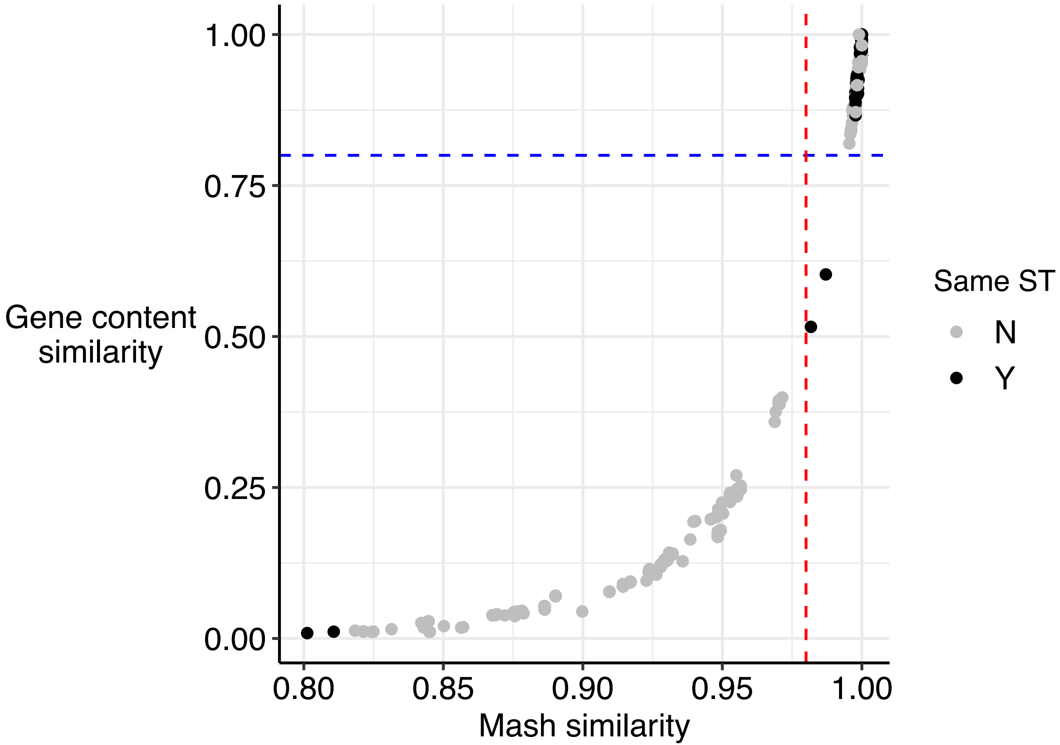

### Supplementary Figure 2

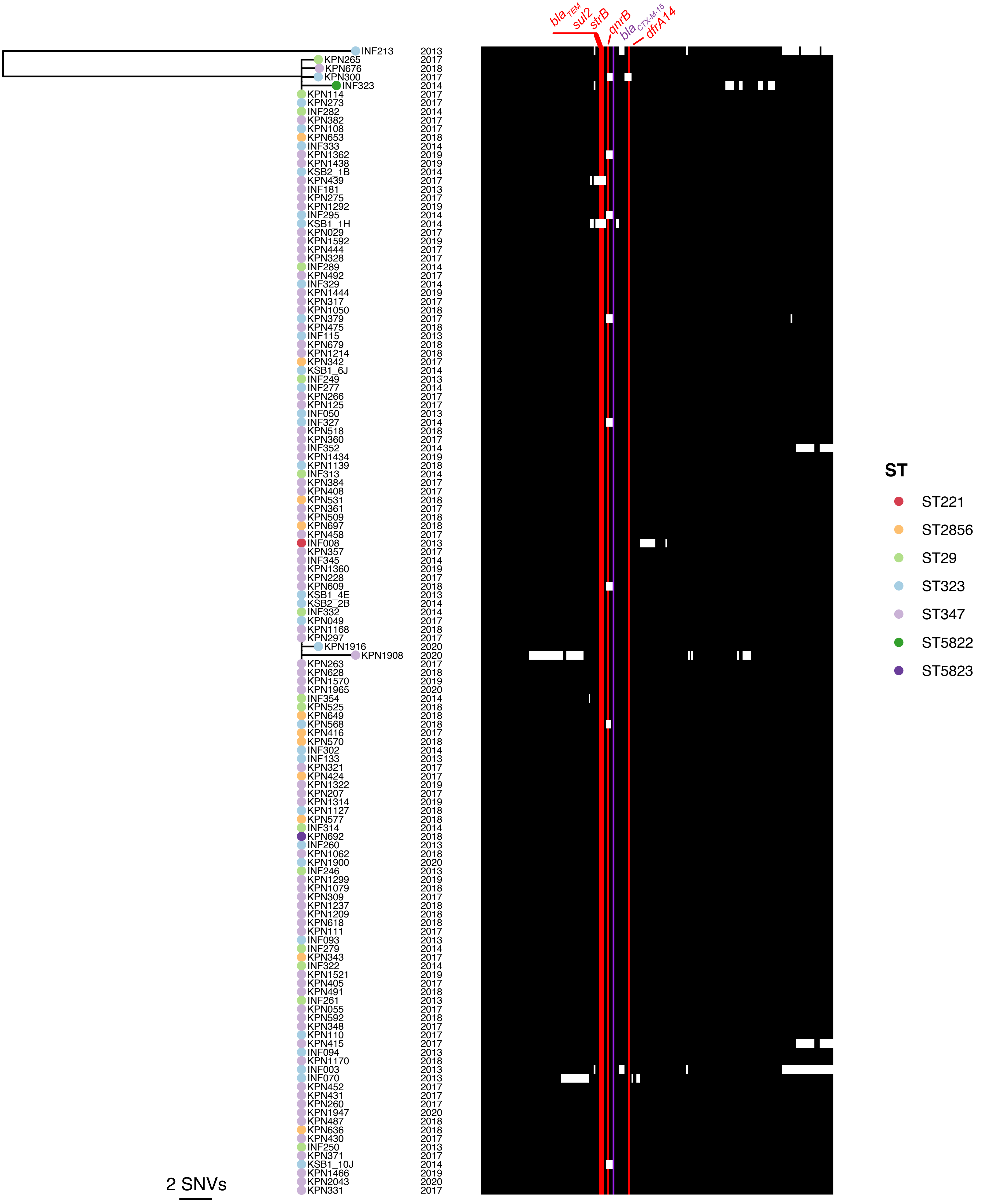

### Supplementary Figure 3

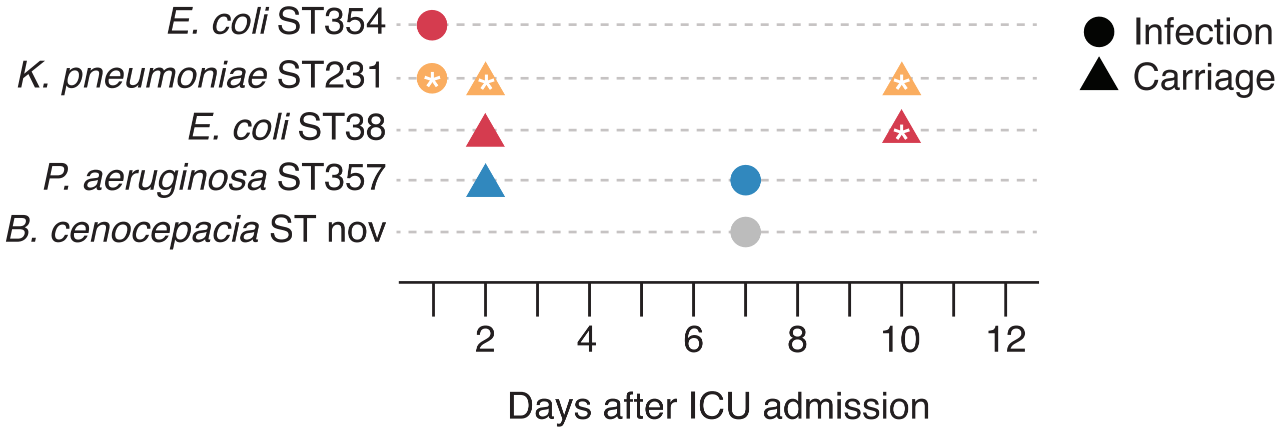

### Supplementary Figure 4

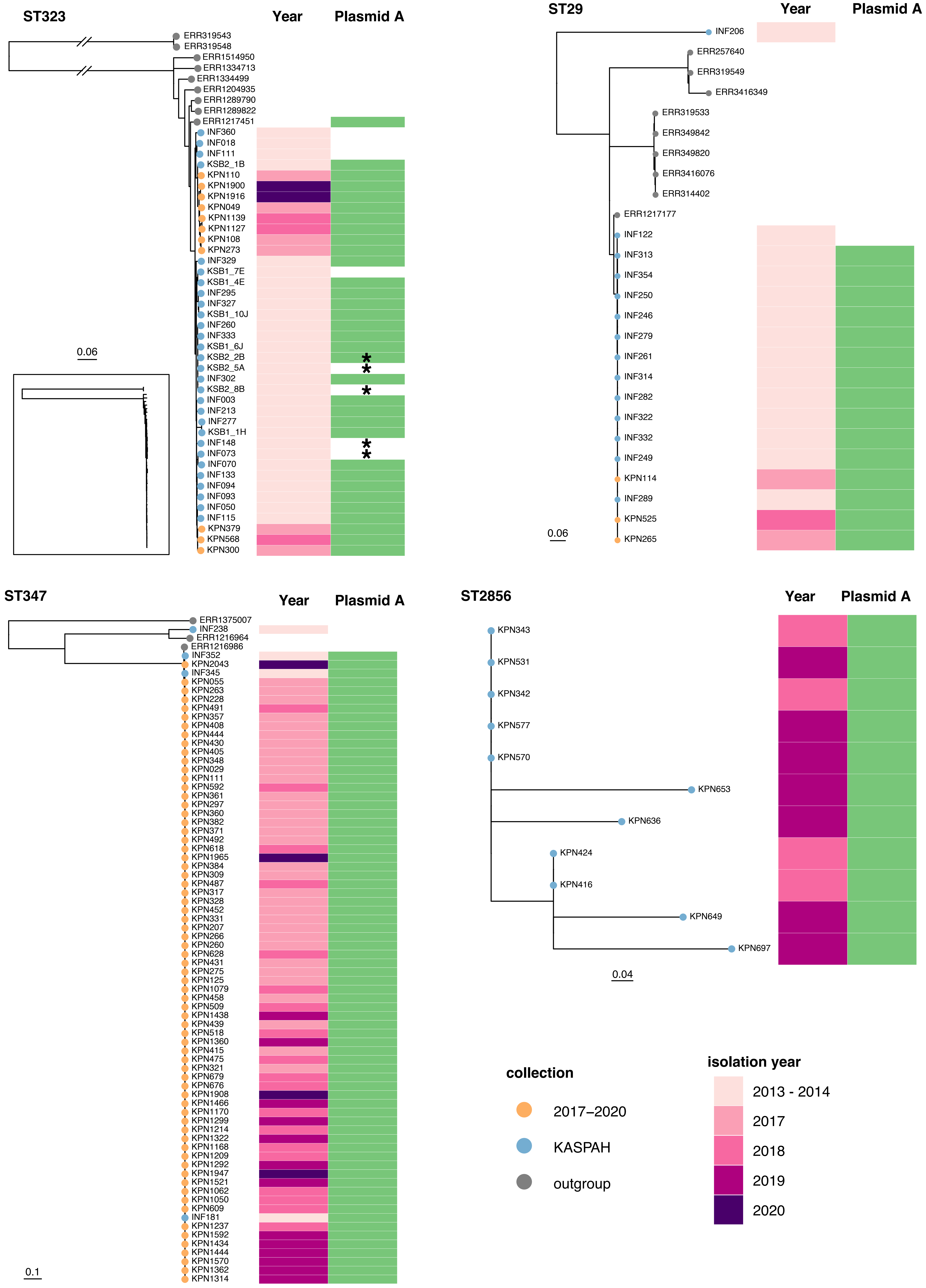

### Supplementary Figure 5

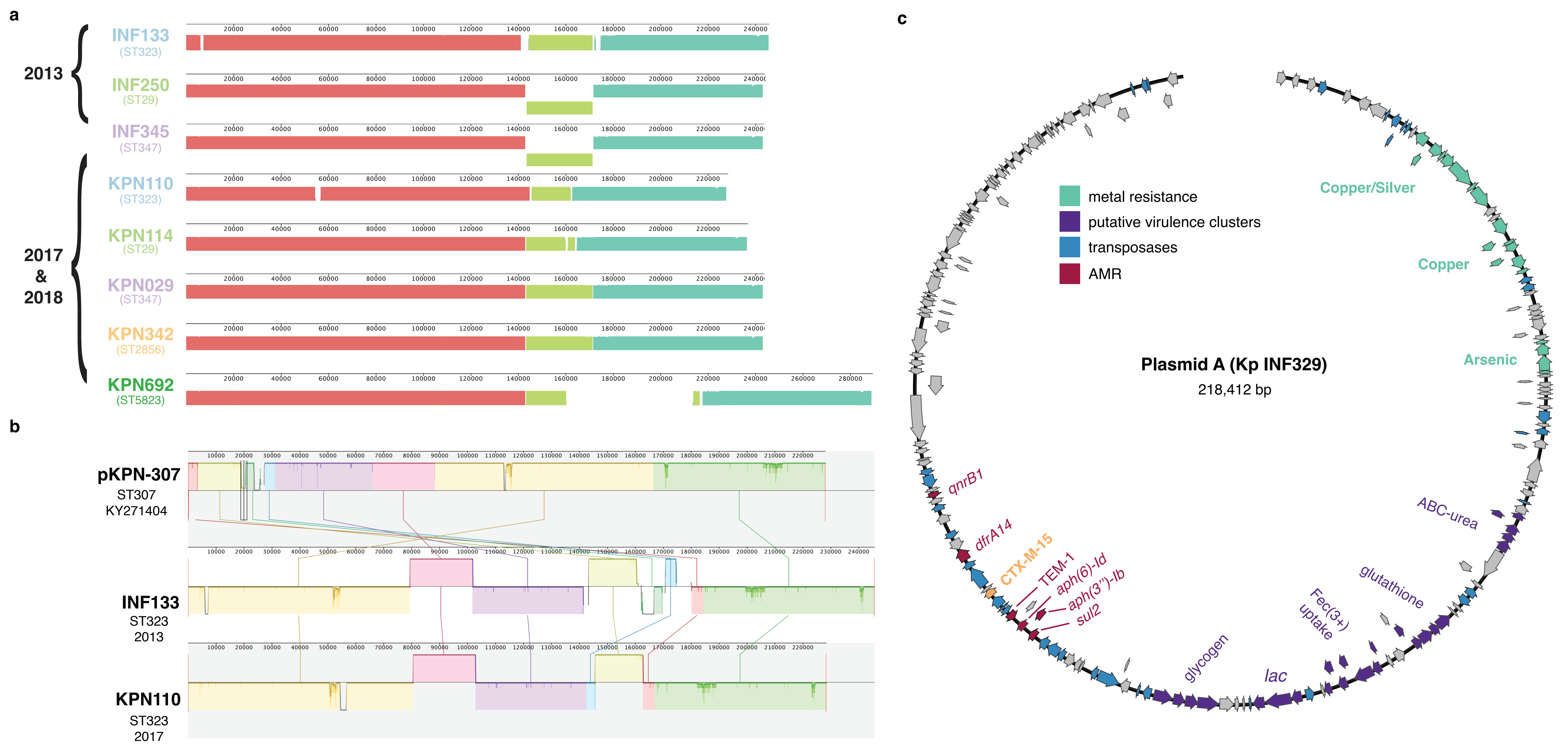
