## Supplementary Text for "ESBL plasmids in *Klebsiella pneumoniae*: diversity, transmission, and contribution to infection burden in the hospital setting"

**Supplementary Methods**

*Phylogenetic analyses*

We generated core-genome phylogenies for *Klebsiella pneumoniae* ST323 (n=40), ST29 (n=17), ST2856 (n=10), and *Klebsiella variicola* ST347 (n=54). For each ST, we included publicly available genomes to outgroup root each tree, with the exception of ST2856 where no publicly available genomes were available (ST323, n=6; ST29, n=9; ST347, n=3; see **Supplementary Table 3** for details of outgroup genomes). Illumina reads were mapped to a completed reference sequence generated from study isolates (INF018 for ST323, accession LR890493; INF250 for ST29, accession LR890374; KPN342 for ST2856, accession CP089384; INF345 for ST347, accession LR890399) using RedDog to identify single nucleotide variants (SNVs) as described previously [1]. The resulting SNV alignments were each filtered to include only SNV sites with allele calls in 100% of genomes. The resulting alignments (ST323 = 7450 SNVs; ST29 = 17,430 SNVs; ST2856 = 13 SNVs; ST347 = 53,093 SNVs) were subjected to maximum likelihood phylogenetic inference using RAxML v8.2.9 [2] with a general time reversible (GTR) substitution model. We generated a Plasmid A phylogeny by mapping all genomes belonging to these STs to the pINF329 reference sequence (accession LR890241) using RedDog as described above. Genomes positive for Plasmid A (n=133) were included in the Plasmid A phylogeny, constructed using RAxML v8.2.9 [2] as outlined above (alignment length 63 SNVs).

**Supplementary Text**

*Carbapenemase plasmids*

The following two STs carried a carbapenemase plasmid in addition to their ESBL plasmid: (i) *K. pneumoniae* ST340 (n=4 isolates), which carried both an IncFIA/IncR ESBL plasmid harbouring *bla*_CTX-M-15_ plus nine other AMR genes and an IncC carbapenemase plasmid harbouring *bla*_IMP-4_ plus five other AMR genes; and (ii) *K. pneumoniae* ST231 (n=8 isolates), which carried an IncC ESBL plasmid harbouring *bla*_CTX-M-15_, *bla*_VEB-1_ and 12 other AMR genes, plus an IncL/M carbapenemase plasmid harbouring *bla*_OXA-48_ without other AMR genes (**Supplementary Table 2**).

The IncL/M carbapenemase plasmid from *K. pneumoniae* ST231 (Plasmid M, harbouring *bla*_OXA-48_, **Table 2**) was found in a burns patient (AH0269) who had three independent *Enterobacteriaceae* carriage episodes plus three independent infections (**Supplementary Figure 3**). This patient was colonised by three unique strains (*K. pneumoniae* ST231 and *E. coli* ST38 detected at two and six days after ICU admission, and *Pseudomonas aeruginosa* ST357 detected at two days after admission), plus one vancomycin resistant *Enterococcus* (ST17, detected six days after admission) [3]. Additionally, *K. pneumoniae* ST231 and *E. coli* ST354 were isolated from wound infections (cultured one day after ICU admission) while *P. aeruginosa* ST357 and *Burkholderia cenocepacia* were isolated from central venous catheter-associated infections (seven days after admission) (**Supplementary Figure 3**). All of the *K. pneumoniae* isolated from patient AH0269 were carbapenem-resistant, harbouring Plasmid M.

The first another *E. coli* ST38 isolate (MSB1_7A) was cultured from the day two rectal swab and was lacking Plasmid M. Subsequently, an *E. coli* ST38 isolate that carried Plasmid M (MSB1_3B) was cultured from the day six rectal swab. High resolution comparison of the two *E. coli* isolates indicated no single nucleotide variants (SNVs) differentiating their chromosomes. Hence we concluded that Plasmid M was likely transferred from a *K. pneumoniae* ST231 donor to an *E. coli* ST38 recipient within the patient’s gut microbiota. The genome data also suggests that the Plasmid M-positive *K. pneumoniae* ST231 strain was transmitted to 4 other patients in the ICU (≤8 core genome SNVs, 0 plasmid SNVs, also discussed previously [3]). Plasmid M is highly similar to the *bla*_OXA-48_ IncL plasmid previously described in *K. pneumoniae* [29] (NC_019154 is 99% identical with 95% coverage to Plasmid M), which has previously been shown to transmit between multiple species within the *Enterobacteriaceae*, including *K. pneumoniae, Enterobacter cloacae, Salmonella enterica* and *E. coli*, most frequently within the gastrointestinal tracts of hospital patients [4].

**Supplementary Legends**

**Supplementary Table 1: Details of all isolates used in this study.**

**Supplementary Table 2: Details of unique ESBL and CP plasmids found in this study.** Kp – *K. pneumoniae*; Kv – *K. variicola*; Ec – *E. coli*; Em – *E. marmotae*.

**Supplementary Table 3: Details of outgroup genomes used for chromosomal ST323, ST29 and ST347 trees.**

**Supplementary Figure 1: Pairwise comparison of *bla*_CTX-M-15_ plasmid sequences by pairwise mash similarity and jaccard gene content similarity.** Each dot represents a pairwise comparison, dots are coloured black if the pair come from the same ST, and grey if they do not. The red dotted line indicates the mash similarity cutoff (0.98), and the blue dotted line the gene content similarity cutoff (0.80). Plasmid sequence pairs whose values were above both cutoffs (upper right corner of the graph) were considered the same plasmid.

**Supplementary Figure 2: Maximum likelihood phylogeny of Plasmid A**. Phylogeny is midpoint rooted. Tips are coloured by ST as per legend. Year of isolation is indicated next to each tip. The heatmap shows the presence of each gene on Plasmid A, with a coloured square indicating presence, and white indicating absence. AMR genes found on Plasmid A are indicated by red boxes, with *bla*_CTX-M-15_ shown in purple.

**Supplementary Figure 3: Timeline for ICU stay of patient AH0269.** Each row indicates a different bacterial lineage colonising this patient, with circles (infection) and triangles (carriage) representing the sampled isolates. White asterisks indicate an isolate carrying Plasmid M encoding the carbapenemase *bla*_OXA-48_.

**Supplementary Figure 4: Maximum-likelihood phylogenies of genomes from the four major STs that carried Plasmid A in KpSC.** ST323, ST29 and ST347 phylogenies are outgroup rooted using publicly available genomes; ST2856 phylogeny is midpoint rooted. Tips of the tree are coloured by collection. Column 1 of the heatmap indicates year of isolation and presence (green) or absence (white) of Plasmid A in column 2. Black stars next to isolates indicate the presence of a chromosomal copy of *bla*_CTX-M-15_.

**Supplementary Figure 5: Comparison of completed Plasmid A sequences. a,** progressiveMauve alignment of completed Plasmid A sequences from one representative of ST323 (blue), ST29 (green) and *K. variicola* ST347 (purple) from 2013, and one representative of ST323, ST29, ST347, ST2856 and ST5823 from 2017 or 2018. **b,** progressiveMauve alignment of the *bla*_CTX-M-15_ plasmid found in *K. pneumoniae* ST307 (accession KY271404) with Plasmid A from ST323 (in both 2013 and 2017). **c,** Gene map of Plasmid A in INF329 (ST323). Genes are represented by arrows and coloured by function as per legend. Putative virulence clusters are the same as those defined by Villa et al.
